# PGAP3 regulates human bronchial epithelial cell mRNAs present in asthma and respiratory virus reference data sets

**DOI:** 10.1101/2024.07.03.24309917

**Authors:** Eric Leslie, Marina Miller, Allison Lafuze, Sofya Svyatskaya, Gil-Soon Choi, David H. Broide

## Abstract

PGAP3 is a glycosylphosphatidylinositol (GPI) phospholipase gene localized within chromosome 17q12-21, a region highly linked to asthma. Although much is known about the function of other chromosome 17q12-21 genes expressed at increased levels in bronchial epithelium such as ORMDL3 and GSDMB, little is known about the function of increased PGAP3 expression in bronchial epithelium in the context of asthma. The aim of this study was therefore to determine whether increased PGAP3 expression in human bronchial epithelial cells regulated expression of mRNA pathways important to the pathogenesis of asthma by utilizing RNA-sequencing and bioinformatic analysis. We performed RNA-sequencing on normal human bronchial epithelial cells transfected with PGAP3 for 24 and 48 hours. PGAP3 regulated genes were compared to asthma and respiratory virus (influenza A, rhinovirus, respiratory syncytial virus) reference data sets to identify PGAP3 target genes and pathways. Approximately 9% of the upregulated PGAP3-induced genes were found in an asthma reference data set, 41% in a rhinovirus reference data set, 33% in an influenza A reference data set, and 3% in a respiratory syncytial virus reference data set. PGAP3 significantly upregulated the expression of several genes associated with the innate immune response and viral signatures of respiratory viruses associated with asthma exacerbations. Two of the highest expressed genes induced by PGAP3 are RSAD2, OASL, and IFN-λ, which are anti-viral genes associated with asthma. PGAP3 also upregulated the antiviral gene BST2, which like PGAP3 is a GPI-anchored protein. We conclude that PGAP3 expression in human bronchial epithelial cells regulates expression of genes known to be linked to asthma, and also regulates the bronchial epithelial expression of genes pertinent to the pathogenesis of respiratory viral triggered asthma exacerbations.

## Introduction

Chromosome 17q12-21 is highly linked to asthma as has been observed in multiple genetic epidemiologic studies^1–3^. The biologic function of two of the nine genes located in chromosome 17q12-21 (ORMDL3 and GSDMB)^2–5^ have been well studied and both genes have been found to promote airway remodeling, which could contribute to the development of asthma. The chr:17q12-21 region contains proximal (PGAP3-ERBB2), core (IKZF3-ZPBP2-GSDMB-ORMDL3), and distal (GSDMA) regions, which have been suggested as independent regions associated with asthma^2^. Studies of the relationship of PGAP3 SNPs and asthma have predominantly focused on three PGAP3 SNPs, namely rs2517<u>954,</u> rs2517<u>955,</u> and rs2941<u>504</u>^6–11^, which are subsequently abbreviated <u>954</u>, <u>955</u>, and <u>504</u>. Several studies have demonstrated that these three PGAP3 SNP risk alleles are associated with increased asthma susceptibility, increased asthma severity, and increased longitudinal asthma exacerbations^6–8^. SNP <u>955</u> in PGAP3 is significantly associated with longitudinal asthma exacerbations after multiple-test adjustment^10^. In addition, two SNPs in PGAP3 (<u>955</u>, <u>504</u>) have risk alleles associated with increased PGAP3 expression^10^. eQTL studies of PGAP3 demonstrated that PGAP3 SNP <u>954</u> has an independent association with increased PGAP3 lung expression^11^, while PGAP3 SNP <u>955</u> is a strong eQTL for PGAP3 in lung tissue^11^. Although there is thus clear evidence from eQTL studies^11^ as well as genetic association studies^6–8^ that the risk alleles of PGAP3 associate with PGAP3 expression, asthma susceptibility, asthma severity, and asthma exacerbations, there are currently no published studies of the biology of PGAP3 in asthma to explain why there is this association, which is the focus of the current study.

PGAP3 (Post-GPI Attachment to Proteins 3) (also known as PERLD1^9^ and PER1^12^) is a chr:17q12-21 localized gene that encodes a glycosylphosphatidylinositol (GPI)-specific phospholipase that primarily localizes to the Golgi apparatus^12^, but can also be detected in the plasma membrane and cytosol (data from Human Protein Atlas v23.proteinatlas.org: https://www.proteinatlas.org/ENSG00000161395-PGAP3)^13^. PGAP3 encodes a seven-transmembrane protein in the Golgi that removes unsaturated fatty acids from the sn-2 position of GlycosylPhosphatidyliInositol (GPI)^12^. GPI anchoring is a posttranslational modification that tethers proteins to plasma membranes, and thus plays a key role in protein sorting and trafficking^12^. There are over 150 human GPI-anchored proteins (GPI-APs), including receptors, adhesion molecules, and enzymes^12^, which underscores the importance of the genes including PGAP3 known to be involved in the biosynthesis and remodeling of the GPI anchor. The GPI anchor is formed in the ER, transported to the Golgi for fatty acid remodeling and cellular export to lipid rafts in the cell membrane^12,13^. The PGAP3 SNP <u>504</u> linked to asthma^9^ has also been associated with peripheral blood mononuclear cell surface and secreted GPI-AP levels^9^, underscoring a potential mechanistic link between PGAP3, GPI-AP, and lipid rafts. Although there are no studies of the function of PGAP3 related to asthma, PGAP3 is known to regulate lipid rafts^14^ that contain GPI-AP, receptors, adhesion molecules, signaling molecules, and enzymes of potential importance to asthma.

As increased expression of other chr 17q12-21 genes (i.e. ORMDL3, GSDMB)^2–5^ in bronchial epithelial cells has identified genes and pathways important to the pathogenesis of asthma, the aim of this study was to determine whether increased PGAP3 expression in normal human bronchial epithelial cells (NHBE) regulated expression of mRNA pathways important to the pathogenesis of asthma by utilizing RNA-sequencing and bioinformatic analysis.

## Methods

### Normal Human Bronchial Epithelial Cell Cultures (NHBE)

NHBE (ATCC) were cultured in 75 mL flasks with 25 mL epithelial-specific media comprised of 98% bronchial epithelial cell media, 1% epithelial cell growth serum, and 1% 10,000 U/mL penicillin (ScienCell). For experiments, NHBE were passaged and subdivided for continued stock culturing (ζ 1x10^6^ cells) or transferred to standard 6-well plates (1x10^6^ per well in 2 mL media) for 24 hours at 37°C prior to experiments. Experiments were completed within 3 – 7 passages.

### PGAP3 Transfection of NHBE

To study the effects on biological pathways of increased PGAP3 expression in NHBE (which do not express significant levels of PGAP3), plasmid PGAP3 (PGAP3) was transfected into NHBE using similar methods as has previously been used to study the biology of increased ORMDL3 or increased GSDMB expression in NHBE^15,16^. Transfection solutions were prepared with an expression vector containing PGAP3 plasmid or with only the expression vector (empty vector) obtained from Origene (PGAP3 Human Untagged Clone, Accession No. NM_033419, vector: pCMV6-XL4, https://www.origene.com), lipofectamine (Invitrogen), and serum-free media (Opti-MEM, Cat No. 11058021, Invitrogen). Solutions were prepared with the following final volumes per well: 1 μL plasmid, 15 μL lipofectamine, and 234 μL DMEM media. After solutions were incubated for 5 minutes, epithelial-specific media was aspirated, cells were washed with 1x PBS (Gibco), and 250 μL transfection solution was added to each well. An additional 750 μL DMEM media was added to each well for a final volume of 1 mL per well. NHBE were transfected for 24 hours and 48 hours prior to RNA-sequencing.

### NHBE RNA Extraction and cDNA Synthesis

After transfection with PGAP3, RNA was extracted from NHBE using a RNeasy Mini Kit (50, Cat No. 74104, Qiagen). Following the manufacturer’s instructions, cells were washed with PBS prior to adding 350 mL lysate buffer (Buffer RLT) to each well. After obtaining RNA diluted in RNase-free water, 2 μL of sample were used to measure RNA quantity and quality using a NanoDrop 2000/2000c spectrophotometer (ND2000, Thermo Fisher Scientific). Samples were then diluted in RNase-free water to standardize concentrations of 1 μg RNA in a final volume of 20 μL, added to cDNA EcoDry Premix (Cat No. 639543, TaKaRa) and reverse transcribed using an Eppendorf Mastercycler (MilliporeSigma) set to 42°C for 60 min and 70°C for 10 min.

### RNA-Sequencing and Statistical Analysis of PGAP3-transfected NHBE

Confirmation of PGAP3 transfection was assessed via quantitative polymerase chain reaction (qPCR, PGAP3 primer Assay ID: Hs.PT.58.19820740, IDT). Three PGAP3-transfected and three control samples were sent to GENEWIZ (Azenta Life Sciences) for RNA-sequencing where differential gene expression was obtained from their automated workflow. In brief, sequence reads were assessed for quality, trimmed using Trimmomatic v.0.36, and mapped to the human reference genome GRCh38 using the STAR aligner v.2.5.2b. Unique gene hit counts were calculated using the Subread package v.1.5.2 and filtered for reads that fell within exon regions. Differential expression analysis was performed using DESeq2. Significance and log2 fold change (log2FC) were determined via the Wald test.

### Bioinformatic Analysis of PGAP3-Regulated Genes and Comparison to Reference Data Sets

We performed bioinformatic and statistical analysis on the following data sets:

1. PGAP3 transfected NHBE vs Empty Vector transfected NHBE (described above).

2. PGAP3 transfected NHBE vs Asthma reference data set

Differentially expressed genes (*p* < 0.05) were analyzed by overrepresentation, gene set enrichment, and network topology-based analysis using WebGestalt (https://www.webgestalt.org) (Liao 2019). For overrepresentation analysis, an asthma reference data set was used that included genes associated with 61 asthma-specific loci discovered by Pividori et al. (2019)^17^ in a genome-wide association study of the UK BioBank data from 37,846 self-reported asthma patients. Specifically, we identified protein-coding genes associated with the 61 asthma loci from a search in National Center for Biotechnology Information (NCBI) Gene Database (https://www.ncbi.nlm.nih.gov/gene/?term=) on March 28, 2023 searching with the 61 asthma loci and then with the advanced search function ‘Default Map Location’ and results filtered for “current” and “protein-coding.” This search identified 2326 protein-coding genes, which we used as our asthma reference data set in our analysis. For network topology analysis, the protein-protein interaction network from BIOGRID was used. Significant Gene Ontology (GO) biological processes of non-redundant genes and Kyoto Encyclopedia of Genes and Genomes (KEGG) pathways were obtained from overrepresentation and gene set enrichment analyses while seed genes were obtained from network topology analysis. Inclusion parameters used for analysis (as applicable) included 5-2000 genes per category, false discovery rate significance level defined as < 0.05, and Benjamin-Hochberg multiple test adjustment.

3. PGAP3 transfected NHBE vs Respiratory Virus reference data sets

Based on bioinformatics analyses, differentially expressed genes were also compared to other RNA-seq data sets regarding NHBE infected with influenza A (Gao et al. 2021)^18^, rhinovirus (Helling et al. 2020)^19^, or respiratory syncytial virus (Mayer et al. 2007)^20^ to compare PGAP3-induced genes with respiratory viruses known to cause exacerbations of asthma. The influenza A virus reference data set acquired from Gao et al. (2021)^18^ contains significantly differentially expressed genes (FDR < 0.05), determined by single-cell RNA-sequencing, of human nasal upper airway epithelial cells collected from adult patients with active influenza A virus infection. While nasal epithelium may not directly reflect the expression of specific genes within bronchial epithelium, nasal samples have previously been studied to determine the effects of 17q21 risk alleles^21^ and in another study the functions of differentially expressed genes in asthmatic bronchial and nasal epithelium were shown to be similar^22^. Therefore, the reference data set from Gao et al. (2021) was included for further analysis regarding any mechanistic changes due to increased PGAP3 expression. The rhinovirus reference data set acquired from Helling et al. (2020)^19^ contains differentially expressed genes (FDR < 0.05), determined by RNA-sequencing, of human asthmatic vs. vehicle-treated bronchial epithelial cells infected with rhinovirus for 24 hours. The respiratory syncytial virus reference data set acquired from Mayer et al. (2007)^17^ contains significantly regulated genes, determined by microarray analysis, of human BEAS-2B cells (an immortalized human bronchial epithelial cell line) infected with respiratory syncytial virus for 24 hours.

### GPI-Anchored Proteins among Significantly Differentially Expressed Genes

Given PGAP3 being a GPI protein and its role in GPI-AP remodeling, GPI-APs were searched for among differentially expressed genes (*p* < 0.05). For this, a reference list of GPI-APs was obtained from UniProt (https://www.uniprot.org)^23^ on August 23, 2023. This list was obtained by searching for “GPI-anchored protein” and filtering for Human, resulting in a reference list of 168 GPI-APs.

### Data Availability Statement

The RNA-sequencing data set generated and analyzed in the current study are available on Figshare: https://doi.org/10.6084/m9.figshare.24666630.v1.

## Results

### Increased PGAP3 upregulated multiple genes in NHBE

In total, 654 genes at 24 hrs and 308 genes at 48 hrs were upregulated (*p* < 0.05) after NHBE were transfected with PGAP3 compared to empty vector control (**Table 1**). Unidentified genes were denoted with “NA” and removed from further analysis.

**Table 1.** PGAP3-Regulated Genes in Normal Human Bronchial Epithelial Cells.

|  | **24 Hours** | **48 Hours** |
| --- | --- | --- |
| **Upregulated** | 654 | 308 |
| **Downregulated** | 581 | 279 |

PGAP3 vs. empty vector expressed genes that achieved significance at an alpha level of 0.05 were included. The data set includes two uncharacterized, upregulated genes that were given a gene symbol of "NA." These genes are included in the totals provided here, but were removed from subsequent analyses.

### Comparison of PGAP3-upregulated genes in NHBE to reference asthma data set

To better understand the relationship between PGAP3-induced differential gene expression to asthma and to respiratory viruses that promote asthma exacerbations and/or the development of asthma (rhinovirus, influenza A, respiratory syncytial virus), PGAP3 upregulated genes were compared to reference data sets. In the UK BioBank asthma reference data set^17^, 62 PGAP3 upregulated genes expressed at 24 hrs were detected, and 36 PGAP3 upregulated genes expressed at 48 hrs were detected (**Table 2**).

**Table 2.** PGAP3-Regulated Genes Found in Reference Data Sets.

**2A. PGAP3-Regulated Genes Found in Asthma Reference Data Sets**
| Reference Data Set | 24 Hours |  | 48 Hours |  |
| --- | --- | --- | --- | --- |
|  | Upregulated | Downregulated | Upregulated | Downregulated |
| UK BioBank Asthma<br>(Pividori et al. 2019,<br>2326 genes) | 62 | 60 | 36 | 27 |

**2B. PGAP3-Regulated Genes Found in Respiratory Virus Reference Data Sets**
| Reference Data Set | 24 Hours |  | 48 Hours |  |
| --- | --- | --- | --- | --- |
|  | Upregulated | Downregulated | Upregulated | Downregulated |
| Influenza A Virus<br>(Gao et al. 2021,<br>3376 genes) | 217 | 70 | 147 | 34 |
| Rhinovirus<br>(Helling et al. 2020,<br>3380 genes) | 267 | 88 | 163 | 38 |
| Respiratory Syncytial<br>Virus (Mayer et al. 2007,<br>492 genes) | 22 | 11 | 9 | 6 |

PGAP3-regulated genes that achieved significance at an alpha level of 0.05 were included.

### Comparison of PGAP3 Upregulated genes in NHBE to reference respiratory virus data sets

In comparing the PGAP3 upregulated genes in NHBE to the respiratory virus reference data sets, a large number of PGAP3 upregulated genes were found in the influenza A reference data set (217 genes at 24 hrs and 147 genes, as well as the rhinovirus reference data set (267 genes at 24 hrs and 163 genes at 48 hrs) (**Table 2**). Relatively fewer PGAP3 regulated genes were also found in the respiratory syncytial virus reference data set (22 genes at 24 hrs and 9 genes at 48 hrs) (**Table 2**).

### Top 10 PGAP3 Upregulated and Top 10 PGAP3 Downregulated genes in NHBE

**Tables 3 and 4** lists the top 10 upregulated and top 10 downregulated differentially expressed (*p* < 0.05) genes in PGAP3 transfected NHBE and provides information regarding in which reference data sets these PGAP3 induced genes were also found. The most notable findings are found in the top 10 PGAP3 upregulated genes where several interferon-stimulating genes (IFIT1, IFI6, IFI27, and IFI44L), RSAD2 (which encodes the antiviral IFN-induced protein viperin^24^, and the chemokine CXCL11 are also found in both the influenza A and rhinovirus reference data sets. This suggests that some of the most upregulated genes induced by PGAP3 are involved with the innate immune response to respiratory viruses with predominately antiviral activity. Other notable genes induced by PGAP3 include OASL, which was detected in three reference data sets (asthma, influenza A, rhinovirus), IFNL1 (also called IFN-λ), a prominent antiviral gene expressed after rhinovirus infection^19^, and BST2, which is the only GPI-AP found in the top 10 upregulated genes at 24 or 48 hours and is also found in the influenza A and rhinovirus reference data sets. Among the most downregulated genes, there are several indications of dysregulated calcium signaling through lower expression of genes involved with calcium binding (PAMR1) and calcium-dependent cell adhesion (CDH15), along with other adhesion molecules (CD36, THY1), and HLA-DQA1, a well-known Class II antigen presenting molecule associated with asthma^25^.

**Table 3.** Top 10 PGAP3-Upregulated Genes in NHBE.

| 24 Hours |  |  |  | 48 Hours |  |  |  |
| --- | --- | --- | --- | --- | --- | --- | --- |
| Rank | Gene | Fold Change (log2) | p-value* | Rank | Gene | Fold Change (log2) | p-value* |
| 1 | ZBP1<br>(Z-DNA Binding Protein 1) | 8.02 | < 0.001 | 1<sup>(2,3)</sup> | IFI6<br>(Interferon Alpha Inducible Protein 6) | 7.60 | < 0.001 |
| 2<sup>(2,3)</sup> | RSAD2<br>(Radical S-Adenosyl Methionine Domain Containing 2) | 7.80 | < 0.001 | 2<sup>(2,3)</sup> | IFI27<br>(Interferon Alpha Inducible Protein 27) | 7.23 | < 0.001 |
| 3<sup>(2,3)</sup> | IFI44L<br>(Interferon Induced Protein 44 Like) | 7.59 | < 0.001 | 3<sup>(2,3)</sup> | IFI44L<br>(Interferon Induced Protein 44 Like) | 7.08 | < 0.001 |
| 4<sup>(2,3)</sup> | CMPK2<br>(Cytidine/Uridine Monophosphate Kinase 2) | 7.56 | < 0.001 | 4 | NRIR<br>(Negative Regulator Of Interferon Response) | 7.08 | < 0.001 |
| 5<sup>(2,3,4)</sup> | IFIT1<br>(Interferon Induced Protein With Tetratricopeptide Repeats 1) | 7.55 | < 0.001 | 5<sup>(2,3)</sup> | BST2<br>(Bone Marrow Stromal Cell Antigen 2) | 7.04 | < 0.001 |
| 6<sup>(1,2,3)</sup> | OASL<br>(2'-5'-Oligoadenylate Synthetase Like) | 7.54 | < 0.001 | 6 | PLVAP<br>(Plasmalemma Vesicle Associated Protein) | 7.03 | < 0.001 |
| 7<sup>(2)</sup> | BCO1<br>(Beta-Carotene Oxygenase 1) | 7.50 | < 0.001 | 7<sup>(2)</sup> | RNU4-2<br>(RNA, U4 Small Nuclear 2) | 7.01 | 0.008 |
| 8<sup>(2,3)</sup> | CXCL11<br>(C-X-C Motif Chemokine Ligand 11) | 7.45 | < 0.001 | 8 | AL445490.1<br>(Uncharacterized Gene) | 6.64 | < 0.001 |
| 9<sup>(3)</sup> | IFNL1<br>(Interferon Lambda 1) | 7.43 | < 0.001 | 9 | GH1<br>(Growth Hormone 1) | 6.54 | < 0.001 |
| 10 | NRIR<br>(Negative Regulator Of Interferon Response) | 7.41 | < 0.001 | 10<sup>(2,3)</sup> | XAF1<br>(XIAP Associated Factor 1) | 6.22 | < 0.001 |
| ** | PGAP3<br>(Post-GPI Attachment To Proteins Phospholipase 3) | 5.55 | < 0.001 | ** | PGAP3<br>(Post-GPI Attachment To Proteins Phospholipase 3) | 4.68 | < 0.001 |

\* = Benjamin-Hochberg adjusted *p*-values for upregulated genes (*p* < 0.05). \*\* PGAP3 expression is presented for confirmation of transfection. Bold text = genes upregulated at both 24 and 48 hours. <sup>1</sup> = genes found in UK Biobank asthma reference data set. <sup>2</sup> = genes found in the influenza A infection reference data set (Gao et al. 2021). <sup>3</sup> = Genes found in the rhinovirus infection reference data set (Helling et al. 2020). <sup>4</sup> = Genes found in the respiratory syncytial virus reference data set (Mayer et al. 2007).

**Table 4.** Top 10 PGAP3-Downregulated Genes in NHBE.

| 24 Hours |  |  |  | 48 Hours |  |  |  |
| --- | --- | --- | --- | --- | --- | --- | --- |
| Rank | Gene | Fold Change (log2) | p-value* | Rank | Gene | Fold Change (log2) | p-value* |
| 1 | **AL118508.1**<br>(Uncharacterized Gene) | **-4.26** | **< 0.001** | 1 | **AC011369.2**<br>(Uncharacterized Gene) | **-3.61** | **< 0.001** |
| 2 | **PAMR1**<br>(Peptidase Domain Containing Associated With Muscle Regeneration 1) | **-3.59** | **< 0.001** | 2 | **EDIL3**<br>(EGF Like Repeats And Discoidin Domains 3) | **-2.71** | **< 0.001** |
| 3 | **CASP14**<br>(Caspase 14) | **-3.58** | **< 0.001** | 3<sup>(2,3)</sup> | **CDH15**<br>(Cadherin 15) | **-2.68** | **< 0.001** |
| 4 | **USH1G**<br>(USH1 Protein Network Component Sans) | **-3.54** | **< 0.001** | 4 | **AK5**<br>(Adenylate Kinase 5) | **-2.61** | **< 0.001** |
| 5<sup>(3)</sup> | **PPARGC1A**<br>(PPARG Coactivator 1 Alpha) | **-3.26** | **< 0.001** | 5<sup>(2)</sup> | **MT1E**<br>(Metallothionein 1E) | **-2.58** | **< 0.001** |
| 6<sup>(3)</sup> | **CD36**<br>(CD36 Molecule) | **-3.22** | **< 0.001** | 6 | **PAMR1**<br>(Peptidase Domain Containing Associated With Muscle Regeneration 1) | **-2.53** | **< 0.001** |
| 7<sup>(1)</sup> | **HLA-DQA1**<br>(Major Histocompatibility Complex, Class II, DQ Alpha 1) | **-3.00** | **< 0.001** | 7<sup>(1)</sup> | **THY1**<br>(Thy-1 Cell Surface Antigen) | **-2.32** | **< 0.001** |
| 8 | **OLFML2A**<br>(Olfactomedin Like 2A) | **-2.91** | **< 0.001** | 8 | **DERL3**<br>(Derlin3) | **-2.31** | **0.001** |
| 9<sup>(2)</sup> | **ZBTB16**<br>(Zinc Finger And BTB Domain Containing 16) | **-2.82** | **< 0.001** | 9 | **CENPA**<br>(Centromere Protein A) | **-2.27** | **< 0.001** |
| 10<sup>(1,3)</sup> | **CXCL14**<br>(C-X-C Motif Chemokine Ligand 14) | **-2.80** | **< 0.001** | 10 | **AL118508.1**<br>(Uncharacterized Gene) | **-2.21** | **< 0.001** |

\* = Benjamin-Hochberg adjusted p-values for PGAP3-downregulated genes ( $p < 0.05$ ). Bold text = PGAP3-downregulated genes ( $p < 0.05$ ) at both 24 and 48 hours. <sup>1</sup> = genes found in UK Biobank asthma reference data set. <sup>2</sup> = genes found in the influenza A infection reference data set (Gao et al. 2021). <sup>3</sup> = Genes found in the rhinovirus infection reference data set (Helling et al. 2020).

### Top PGAP3 Upregulated genes found in the asthma and respiratory viral reference data sets

Given a greater number of genes differentially expressed by PGAP3 vs empty vector control at 24 hours and an upregulation of the most significant pathways (discussed in the bioinformatics analysis results below), a closer analysis of PGAP3-induced upregulated genes at 24 hours was used to find connections with the asthma and respiratory viral reference data sets. **Tables 5-7** provide the list of top PGAP3-induced upregulated genes (*p* < 0.05) found in the asthma and respiratory viral reference data sets. These tables highlight several additional genes commonly associated with asthma. PGAP3 regulated genes found in the UK BioBank asthma reference data set include the chemokine CCL5 (also known as RANTES), the STAT inhibitor SOCS1, and the IL-15 receptor IL15RA (**Table 5**). Several additional PGAP3 regulated antiviral genes are shared between the influenza A (**Table 6**) and rhinovirus (**Table 7**) reference data sets, including MX1, MX2, OAS1, and OAS2. The respiratory syncytial virus reference data set shares the following four PGAP3 regulated genes with the other data sets of potential importance to asthma: DDX58 (also known as RIG-I, found in the rhinovirus and influenza A reference data sets), CXCL3 (found in the influenza A reference data set), SOCS3 (found in the influenza A reference data set), and IL6 (found in the asthma and rhinovirus reference data sets) (**Table 8**).

**Table 5.** PGAP3-Upregulated Genes Found in the UK BioBank Asthma Reference Data Set.

| Gene | PGAP3-Induced Fold Change (log2) |
| --- | --- |
| OASL | 7.54 |
| **CCL5 (RANTES)** | **6.88** |
| IFITM1 | 6.07 |
| PGAP3 | 5.55 |
| SLC15A3 | 4.91 |
| IRF7 | 4.90 |
| BATF2 | 4.72 |
| PSMB9 | 4.07 |
| CFB | 3.99 |
| SP8 | 3.55 |
| HLA-F | 3.23 |
| DHX58 | 2.82 |
| IL7 | 2.79 |
| HLA-B | 2.77 |
| IFITM2 | 2.69 |
| IFITM3 | 2.65 |
| HCAR2 | 2.59 |
| SPRR2D | 2.47 |
| NFASC | 2.44 |
| TAP1 | 2.37 |
| HLA-C | 2.26 |
| S100A7 | 2.15 |
| LCE3D | 2.12 |
| HCAR3 | 2.10 |
| **SOCS1** | **1.98** |
| CRCT1 | 1.97 |
| TAP2 | 1.96 |
| HSPB9 | 1.96 |
| **IL15RA** | **1.90** |
| PSMB8 | 1.89 |
| CTSS | 1.82 |
| TMEM229B | 1.81 |
| IRF1 | 1.76 |
| LCE3E | 1.71 |
| SPRR3 | 1.70 |
| ADAR | 1.68 |
| IL18R1 | 1.67 |
| STAT2 | 1.66 |
| CBR3 | 1.64 |
| C6orf15 | 1.63 |
| IL1RL1 | 1.45 |
| TNIP3 | 1.44 |
| HLA-A | 1.41 |
| SLFN5 | 1.39 |
| SP6 | 1.36 |
| TLR1 | 1.29 |
| MAJIN | 1.28 |
| RMI2 | 1.27 |
| PDCD1LG2 | 1.25 |
| HNF4G | 1.24 |
| IL24 | 1.23 |
| PSORS1C1 | 1.22 |
| HUS1B | 1.21 |
| ZFHX4 | 1.20 |
| CYSRT1 | 1.15 |
| RAPGEF5 | 1.12 |
| TMEM171 | 1.11 |
| IL6 | 1.09 |
| STEAP1B | 1.09 |
| ZFYVE26 | 1.09 |
| GAST | 1.01 |
| HLA-E | 1.00 |

PGAP3-upregulated genes that achieved significance at an alpha level of 0.05 were included. Bold text = select PGAP3-upregulated genes associated with asthma that are discussed in text.

**Table 6.** Top 25 PGAP3-Upregulated Genes Found in Influenza A Reference Data Set.

| Gene | PGAP3-Induced Fold Change (log2) |
| --- | --- |
| **RSAD2** | **7.80** |
| **IFI44L** | **7.59** |
| CMPK2 | 7.56 |
| IFIT1 | 7.55 |
| **OASL** | **7.54** |
| BCO1 | 7.50 |
| **CXCL11** | **7.45** |
| **MX2** | **7.40** |
| **MX1** | **7.23** |
| IFI6 | 7.19 |
| BST2 | 7.14 |
| XAF1 | 7.04 |
| ISG15 | 6.99 |
| **CXCL10** | **6.94** |
| **CCL5 (RANTES)** | **6.88** |
| BISPR | 6.87 |
| IFIT2 | 6.86 |
| IFI44 | 6.75 |
| IFIT3 | 6.71 |
| HERC5 | 6.64 |
| GBP4 | 6.53 |
| IDO1 | 6.49 |
| LAMP3 | 6.43 |
| IFI27 | 6.41 |
| MMP13 | 6.33 |

PGAP3-upregulated genes that achieved significance at an alpha level of 0.05 were included. Bold text = PGAP3-upregulated genes associated with asthma that are discussed in text.

**Table 7.** Top 25 PGAP3-Upregulated Genes Found in Rhinovirus Reference Data Set.

| Gene | PGAP3-Induced Fold Change (log2) |
| --- | --- |
| ZBP1 | 8.02 |
| **RSAD2** | **7.80** |
| **IFI44L** | **7.59** |
| CMPK2 | 7.56 |
| IFIT1 | 7.55 |
| **OASL** | **7.54** |
| **CXCL11** | **7.45** |
| **IFNL1** | **7.43** |
| **MX2** | **7.40** |
| **MX1** | **7.23** |
| IFI6 | 7.19 |
| BST2 | 7.14 |
| XAF1 | 7.04 |
| ISG15 | 6.99 |
| RASGRP3 | 6.95 |
| **CXCL10** | **6.94** |
| CXCL9 | 6.87 |
| IFIT2 | 6.86 |
| APOBEC3A | 6.81 |
| IFI44 | 6.75 |
| IFIT3 | 6.71 |
| IFNB1 | 6.68 |
| HERC5 | 6.64 |
| GBP4 | 6.53 |
| IDO1 | 6.49 |

**Table 8.** PGAP3-Upregulated Genes Found in Respiratory Syncytial Virus Reference Data Set.

| **Gene** | **PGAP3-Induced Fold Change (log2)** |
| --- | --- |
| IFIT1 | 7.55 |
| IFI44 | 6.75 |
| **DDX58** | **5.24** |
| PMAIP1 | 3.35 |
| HSPA6 | 2.91 |
| **CXCL3** | **2.15** |
| STX11 | 2.08 |
| NAV3 | 1.82 |
| IRF1 | 1.76 |
| **SOCS3** | **1.75** |
| FAM46A | 1.72 |
| BIRC3 | 1.62 |
| ARC | 1.62 |
| FAM46C | 1.24 |
| FOS | 1.24 |
| PLAUR | 1.22 |
| ZFHX4 | 1.20 |
| SOD2 | 1.19 |
| ASPM | 1.15 |
| TRIM38 | 1.11 |
| **IL6** | **1.09** |
| RGS2 | 1.06 |

PGAP3-upregulated genes that achieved significance at an alpha level of 0.05 were included. Bold text = PGAP3-upregulated genes associated with asthma that are discussed in text.

### PGAP3 upregulated genes found in more than one respiratory viral reference data set

**Table 9** provides the top 25 PGAP3 upregulated genes also found in the influenza A, rhinovirus, and respiratory syncytial virus reference data sets. In addition, **Figure 1** provides a diagram to illustrate the overlap of PGAP3-differentially expressed genes found in the three individual respiratory viral reference data sets (rhinovirus, influenza A, respiratory syncytial virus). When analyzing which genes are unique or shared between the data sets, the largest proportion consists of shared genes between the rhinovirus and influenza A reference data sets (43%) while < 2% of the genes were shared between rhinovirus and respiratory syncytial virus reference data sets and between influenza A and respiratory syncytial virus reference data sets. The rhinovirus reference data set had the largest proportion of unique genes (32%), followed by influenza A (18%) and respiratory syncytial virus (< 2%). Seven genes (< 2%) were shared between all three respiratory virus reference data sets: IFIT1, IFI44, DDX58, PMAIP1, IRF1, FAM46A, and PLAUR.

**Figure 1.**
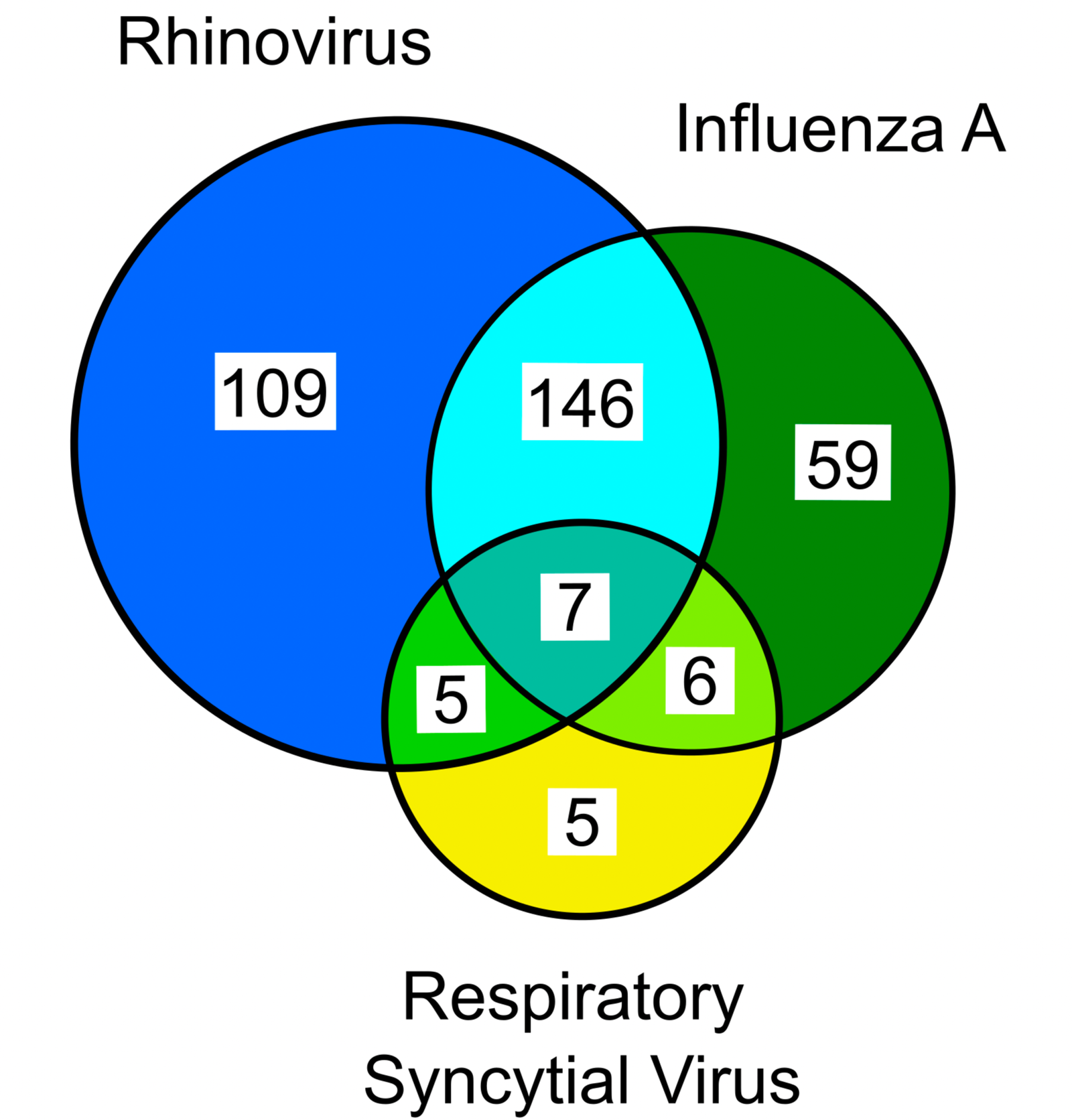
Venn diagram of unique and shared PGAP3 regulated genes found in the three reference data sets of human bronchial epithelial cells infected with respiratory viruses known to cause exacerbations of asthma: rhinovirus, influenza A, and respiratory syncytial virus.

**Table 9.** Top 25 PGAP3-Upregulated Genes Found in Both Influenza A and Rhinovirus Reference Data Sets.

| Gene | PGAP3-Induced Fold Change (log2) |
| --- | --- |
| **RSAD2** | **7.80** |
| **IFI44L** | **7.59** |
| CMPK2 | 7.56 |
| <u>IFIT1</u> | <u>7.55</u> |
| **OASL** | **7.54** |
| **CXCL11** | **7.45** |
| **MX2** | **7.40** |
| **MX1** | **7.23** |
| IFI6 | 7.19 |
| BST2 | 7.14 |
| XAF1 | 7.04 |
| ISG15 | 6.99 |
| **CXCL10** | **6.94** |
| IFIT2 | 6.86 |
| <u>IFI44</u> | <u>6.75</u> |
| IFIT3 | 6.71 |
| HERC5 | 6.64 |
| GBP4 | 6.53 |
| IDO1 | 6.49 |
| LAMP3 | 6.43 |
| IFI27 | 6.41 |
| **OAS1** | **6.31** |
| **OAS2** | **6.07** |
| IFITM1 | 6.07 |
| GMPR | 5.93 |

PGAP3-upregulated genes that achieved significance at an alpha level of 0.05 were included. All 25 PGAP3-upregulated genes were found in both influenza A and rhinovirus reference data sets. Bold text = PGAP3-upregulated genes associated with asthma that are discussed in text. Underline = PGAP3-upregulated genes found in respiratory syncytial virus reference data set. There were 7 PGAP3-upregulated genes found in all three respiratory viral data sets when considering a log2 fold change $\geq 1.0$ : IFIT1, IFI44, DDX58, PMAIP1, IRF1, FAM46A, and PLAUR.

### PGAP3 regulation of GPI-Anchored Proteins

**Table 10** focuses only on the 13 GPI-anchored proteins within the RNA-Seq data set (Table 10A; 6 upregulated: Table 10B; 7 downregulated). There were 6 PGAP3 upregulated GPI-anchored proteins (*p* < 0.05) at 24 hours and 4 PGAP3 upregulated GPI-anchored proteins (*p* < 0.05) at 48 hours. The upregulated GPI-anchored proteins BST2 (7.14 log2 fold change) and PGAP3 had noticeably higher fold changes compared to the other GPI-anchored proteins. All 6 GPI-anchored protein genes upregulated by PGAP3 were found in at least one reference data set (1 in the asthma reference data set, 4 in the influenza A virus reference data set, 5 in the rhinovirus reference data set, and 1 in the respiratory syncytial virus reference data set). The upregulation of the GPI-anchored protein LY6E at both 24 and 48 hours is also notable given its role in enhancing influenza A infection and other viruses, via entry into the cell, in an evolutionarily conserved manner^26^. Overall, these results demonstrate that PGAP3 influenced expression of a subset GPI-APs, in particular BST2 and LY6E.

**Table 10.**
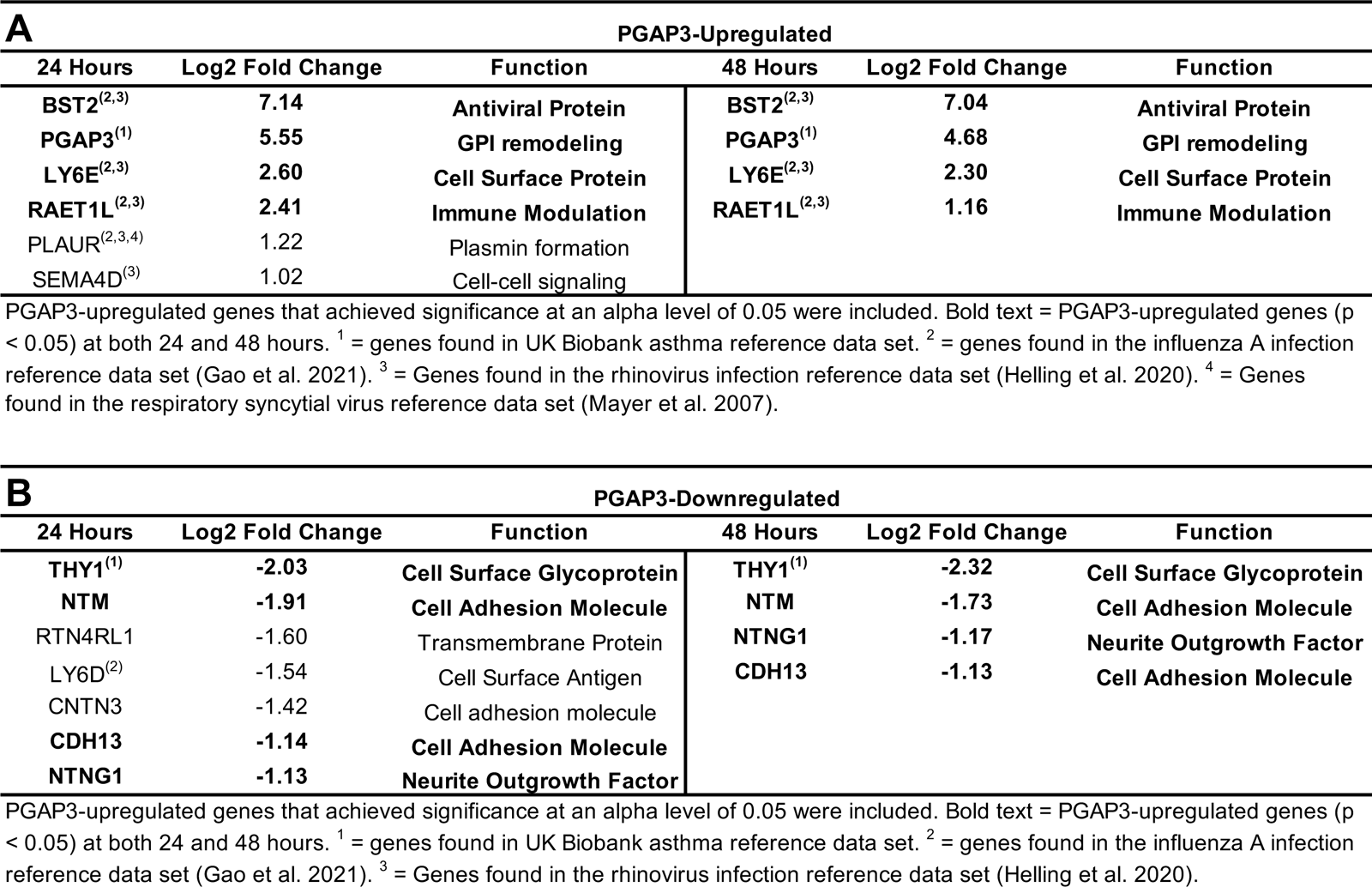
PGAP3-Upregulated GPI-Anchored Proteins.

### PGAP3 regulated pathway analysis by GSEA

Results of the gene set enrichment analysis (GSEA) include 27 positively regulated and 0 negatively regulated GO biological processes at 24 hours while there were 14 positively regulated and 5 negatively regulated GO biological processes at 48 hours. Regarding KEGG pathways, there were multiple positively regulated pathways (9 at 24 hrs; 8 at 48 hours), with no negatively regulated pathways. The top 10 GO biological processes and KEGG pathways based on enrichment score are provided in **Figure 2**. Significant KEGG pathways (**Figure 2A**) generally demonstrated an upregulation of innate immune responses in defense against viral RNA and peptide fragments of infected cells (Cytosolic DNA-sensing as well as RIG-I, NOD-like, and Toll-like receptor signaling) as well as collective changes in gene expression common to several viruses. A key viral pathway detected is influenza A, which is known to promote wheezing in asthma patients or those predisposed to develop asthma^27^. The five downregulated pathways are all involved in the cell cycle (cell cycle G2/M phase transition, regulation of cell cycle phase transition, cytokinesis, mitotic cell cycle phase transition, and chromosome segregation). The GO biological process results (**Figure 2B**) are consistent with the significant KEGG pathways by their associations with innate immunity that includes, but is not limited to, type 1 interferon and cellular defense responses. When the UK BioBank asthma reference data set was used as a background in the overrepresentation analysis (**Figure 3**), the top KEGG pathways (**Figure 3A**) and GO biological processes (**Figure 3B**) are overall consistent with the GSEA since many are involved with viral signatures and the innate immune system.

**Figure 2.**
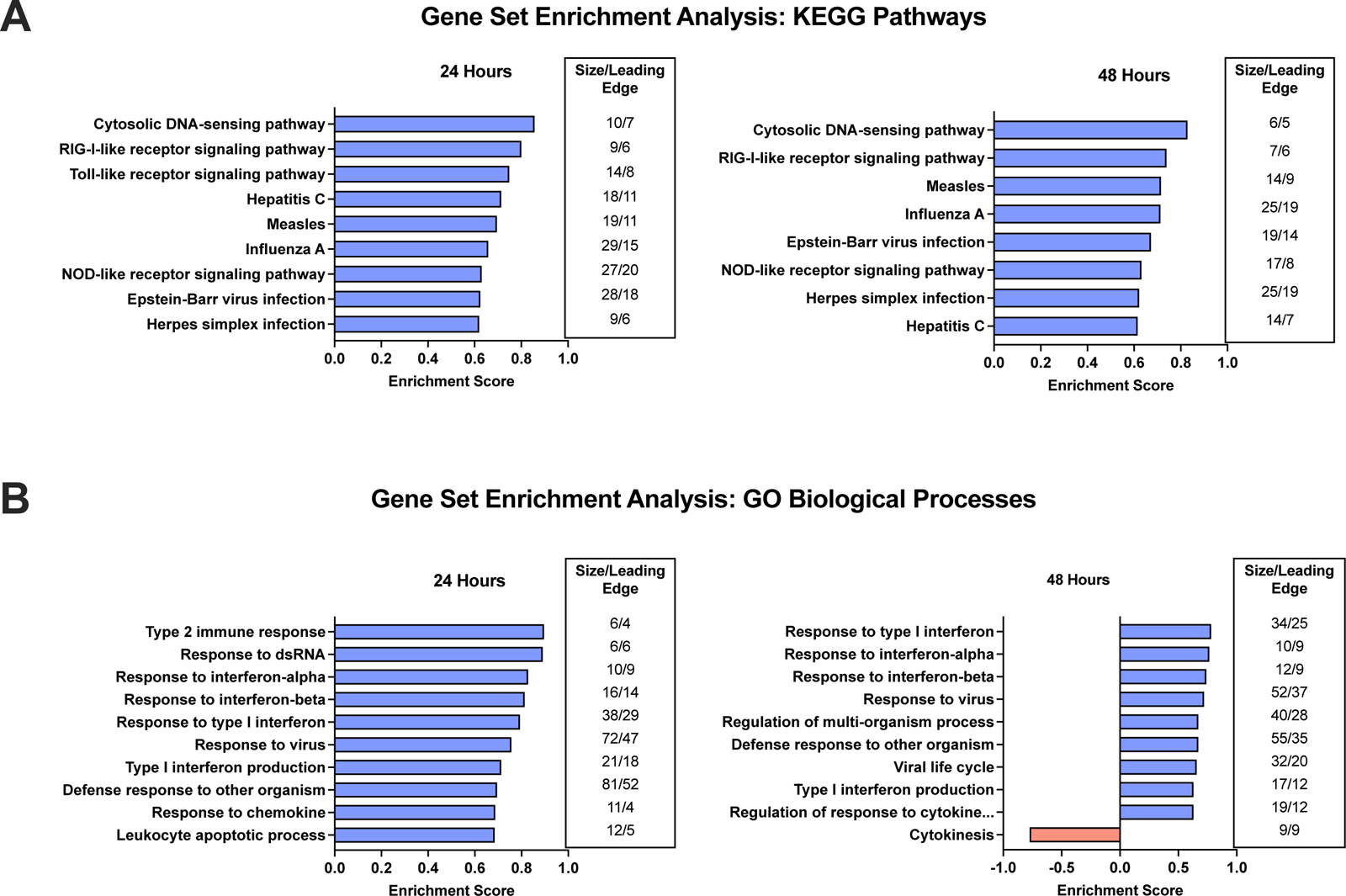
Gene Set Enrichment Analysis. Illustrated are the top 10 KEGG pathways (**2A**) and Gene Ontology (GO) biological processes (**2B**) by enrichment score in human bronchial epithelial cells transfected with PGAP3 for 24 and 48 hours.

**Figure 3.**
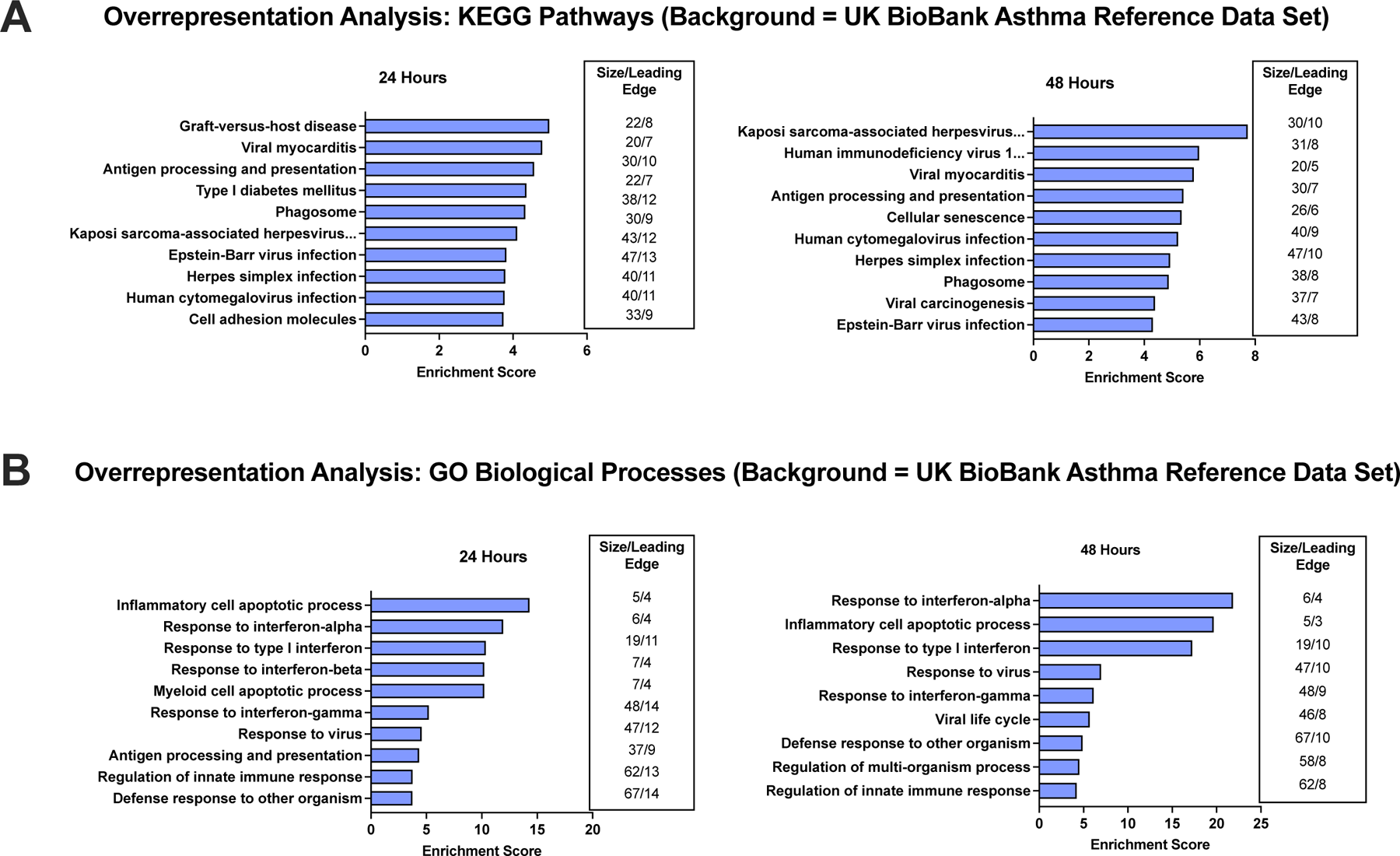
Overrepresentation Analysis. Illustrated are the top 10 KEGG pathways (**3A**) and Gene Ontology (GO) biological processes (**3B**) by enrichment score in human bronchial epithelial cells transfected with PGAP3 for 24 and 48 hours. The UK BioBank asthma reference data set was used, which included 2,326 genes associated with 61 asthma-specific loci identified by Pividori et al. (2019)^17^.

### PGAP3 regulated Pathway Analysis by Highlighted Seeds and Network Topology Analysis

Highlighted seeds with a log2FC ≥ 2.0 were included to identify the most connected or potentially influential seeds (genes) from the network topology analysis (**Table 11**). TRIM25, ISG15, and HLA-C were three of the four highlighted seeds that met these criteria. They are potentially involved in novel pathways that we identified that PGAP3 regulates including RIG-1 signaling and antigen presentation (TRIM25 and ISG15 are involved with RIG-I signaling and HLA-C is involved with antigen presentation).

**Table 11.** Network Topology Analysis Highlighted Seeds.

| **A** |  |  |  |  |
| --- | --- | --- | --- | --- |
| 24 Hours |  |  |  |  |
| Highlighted Seeds | Random Walk Probability | Gene Interactions | Log2 Fold Change | Function |
| **TRIM25<sup>(3)</sup>** | **4.09E-03** | **58** | **2.17** | **Ubiquitin ligase (RIGI, ISG15)** |
| CCNA1<sup>(2,3)</sup> | 9.42E-04 | 2 | 2.21 | Cell Cycle |

| **B** |  |  |  |  |
| --- | --- | --- | --- | --- |
| 48 Hours |  |  |  |  |
| Highlighted Seeds | Random Walk Probability | Gene Interactions | Log2 Fold Change | Function |
| **ISG15<sup>(2,3)</sup>** | **1.61E-03** | **17** | **5.79** | **Chemoattractant; RIG-I activation** |
| **HLA-C<sup>(1,2,3)</sup>** | **1.54E-03** | **10** | **2.67** | **Antigen presentation** |

PGAP3-upregulated genes that achieved significance at an alpha level of 0.05 and had a log2 fold change $\geq 2.0$ were included. Bold text = PGAP3-upregulated genes at both 24 and 48 hours. <sup>1</sup> = genes found in UK Biobank asthma reference data set. <sup>2</sup> = genes found in the influenza A infection reference data set (Gao et al. 2021). <sup>3</sup> = genes found in the rhinovirus infection reference data set (Helling et al. 2020).

## Discussion

As asthma is a disease of the airways the bronchial epithelium plays an important role in the pathogenesis of asthma as inhaled allergens and viruses can trigger the airway epithelium to express pathways important to the pathogenesis of asthma^28^. Increased expression of chromosome 17q12-21 genes such as ORMDL3 and GSDMB have been detected in the bronchial epithelium in asthma and are associated with expression of pathways pertinent to the pathogenesis of asthma^16,29^. In this study, we have identified that increased expression of another chromosome 17q12-21 gene, namely PGAP3, upregulates expression of 654 genes in bronchial epithelium. We compared the genes induced by PGAP3 to an asthma reference data set based on asthma-specific loci discovered from the UK BioBank to determine whether any of the genes induced by PGAP3 were also present in this asthma data set. Approximately 9% of the upregulated genes induced by PGAP3 in NHBE (62 out of 654 genes upregulated by PGAP3) were identified in the asthma reference data set, which further supports PGAP3’s link to asthma. Interestingly, as respiratory viruses are also linked to asthma exacerbations and/or the development of asthma we identified that approximately 41% of the upregulated PGAP3-induced genes in epithelium were found in a rhinovirus reference data set, 33% in an influenza A reference data set, and 3% in a respiratory syncytial virus reference data set. PGAP3 significantly upregulated the expression of several genes associated with the innate immune response and viral signatures of respiratory viruses associated with asthma exacerbations. Two of the highest expressed genes induced by PGAP3 are RSAD2^30^ and OASL^31^, which are anti-viral genes associated with asthma. PGAP3 also upregulated the antiviral gene BST2^32^, which like PGAP3 is a GPI-anchored protein. These studies suggest that PGAP3 expression in human bronchial epithelial cells regulates expression of genes known to be linked to asthma, and that PGAP3 also regulates the bronchial epithelial responses to viruses pertinent to the pathogenesis of respiratory viral triggered asthma exacerbations. Interestingly, the pathways induced by PGAP3 in bronchial epithelium have differences from those induced by other chromosome 17q12-21 genes linked to asthma such as ORMDL3 or GSDMB, but share with ORMDL3 and GSDMB the ability to induce a few selected genes such as OAS^4,16^.

The 62 genes upregulated by PGAP3 we identified as also being present in the UK BioBank asthma reference data set (**Table 5**), include the chemokine CCL5 (also known as RANTES), the STAT inhibitor SOCS1, the IL-15 receptor IL15RA. CCL5 is localized to the same chromosome loci as PGAP3 (17q12-21) and the CCL5 -403G/A polymorphism has been linked to asthma^33^. Recently, high mRNA expression of CCL5 has been correlated with the Th1 cytokines CXCL9 and CXCL10 (also known as IP-10) in sputum from patients with asthma^34^. In mouse models of asthma there is a link between CCL5 and tissue-resident memory T-cell activation as inhibition of CCR5 (the CCL5 receptor) blunts the effect^34^. Several chemokines in addition to CCL5 were upregulated by PGAP3 including CXCL9, and CXCL10, which may indicate expression of PGAP3 promotes recruitment of inflammatory cells into the airway. Another PGAP3 upregulated gene of note is IL15RA, which is a receptor that binds IL-15 and is expressed in the sputum of patients with asthma^35^. IL-15 has also been shown to promote Th2 differentiation and allergic sensitization in a mouse model of asthma^36^. Finally, induction of SOCS1 by PGAP3 may also be important to asthma as RNA-sequencing analysis of human nasal epithelial cells from children with asthma found significant expression of SOCS1^37^ that is known to regulate cytokine signaling in both human bronchial epithelial cells from patients with asthma^38^ and in the lungs of a mouse model of asthma^39^.

Respiratory viruses are important triggers of asthma exacerbations^40^. In addition, RV has been shown to increase the risk of the development of childhood asthma in subjects with a SNP linked to chromosome 17q12-21^41^. Our current study demonstrates that increased expression of PGAP3 in NHBE through induction of antiviral pathways could potentially influence the duration and/or severity of viral infection in epithelium through several mechanisms. For example, PGAP3 expression in NHBE induced robust expression of antiviral genes commonly associated with asthma (RSAD2, OASL, OAS1, OAS2, OAS3, MX1, and MX2), and anti-viral pathways (Cytosolic DNA sensing, RIG-I signaling, NOD-like receptor signaling, and Toll-like receptor signaling). Several other genes including RSAD2, MX1, and BST2 along with ISG15 as highlighted by the network topology analysis have been previously linked to anti-viral responses in epithelial cells ^42^. Induction of these genes and innate pathways by PGAP3 could all potentially limit the extent of a viral infection in epithelium. The connections from the RNA-Seq analysis to RIG-I signaling is particularly notable. RIG-I signaling is known to induce the antiviral genes OASL and the GPI-AP gene BST2^43,44^, which are two of the highest expressed upregulated genes in the PGAP3 data set. OASL is also known to further increase RIG-I signaling, eliciting a positive feedback loop, to improve the antiviral response^31^. While not found in the pathway analysis, IFN-λ was one of the top 10 upregulated genes (FC = 7.43) and has been shown to be positively associated with peak levels of RV infection, despite asthmatic epithelial cells showing a delayed IFN-λ response to rhinovirus^45^. However, since a delayed IFN-λ response to rhinovirus is not always shown^46,47^, whether PGAP3 induces IFN-λ in response, or in this case an anticipated response, to viral infection requires further investigation. The ability of PGAP3 to induce BST2 expression, along with other anti-viral genes, suggests PGAP3 regulates antiviral responses in NHBE. Interestingly, TRIM25, a key RIG-I activator^48^, had the most gene interactions in the network topology analysis, while RIG-I signaling is the second most enriched KEGG pathway. In addition to these PGAP3 regulated anti-viral responses, a smaller number of PGAP3 regulated pro-viral genes were also detected, including LY6E. Given that LY6E enhances cellular infection of influenza A virus among several others (Chikungunya virus, dengue virus, human immunodeficiency virus, West Nile virus, and yellow fever virus)^26^, PGAP3 may contribute to increased susceptibility to viral infection at the same time it coordinates cellular antiviral responses to protect against viruses.

The enriched GO biological processes and KEGG pathways obtained from the PGAP3 overrepresentation analysis, which incorporated the UK BioBank asthma reference data set, largely reflect those found in the gene set enrichment analysis. This suggests that many of the cellular effects of PGAP3 in NHBE include genes from asthma-specific loci. The presence of TRIM25 and ISG15 among the highlighted seeds (FC = 2.0+) further demonstrates PGAP3’s potential connection to antiviral responses via RIG-I signaling. The bioinformatic analyses provided supporting evidence linking PGAP3 overexpression in NHBE to asthma and respiratory viral signatures.

Although we have identified that PGAP3 upregulated 654 genes of which 9% have been identified in an asthma reference data set, and that PGAP3 upregulated genes and pathways linked to innate immunity and antiviral response, there are limitations to the study in terms of the need for future investigations to examine the biological consequences of PGAP3-induced upregulated genes and pathways, which are currently only inferred from the RNA-Seq and bioinformatic analysis. Studies of bronchial epithelial cells from asthmatics with SNPs linked to increased PGAP3 expression would also be of interest as compared to studies of NHBE. However, recruitment of asthmatics with SNPs linked to increased PGAP3 for an invasive bronchoscopy would be needed.

In summary, we have identified that increased expression of the Chr17q12-21 gene PGAP3 in bronchial epithelium upregulates expression of 654 genes in bronchial epithelium approximately 9% of which genes were identified in the UK Biobank asthma reference data set, supporting PGAP3’s link to asthma. The PGAP3 regulated genes detected in the UK Biobank asthma reference data set include RANTES (CCL5), and OASL. Interestingly, we also identified that PGAP3 regulated genes were identified in data sets of respiratory viruses linked to asthma (rhinovirus and influenza A). Two of the highest expressed genes induced by PGAP3 are RSAD2 and OASL, which are antiviral genes associated with asthma^30,31^. PGAP3 also upregulated the antiviral gene BST2, which like PGAP3 is a GPI-anchored protein. These studies suggest that PGAP3 expression in human bronchial epithelial cells regulates expression of genes known to be linked to asthma, and that PGAP3 also regulates the bronchial epithelial responses to viruses pertinent to the pathogenesis of respiratory viral triggered asthma exacerbations. These studies underscore the potential importance of PGAP3 to the biology of asthma.

## Author Contributions

Conception or design of the work (DHB, EL, MM), acquisition (EL, MM, AL, SS, GSC), analysis or interpretation of the work (EL, MM, DB), drafting or revising manuscript for submission (EL, DB).

## Ethical Approval

No ethical approval was obtained because this study does not use human subjects or animals.

## Informed Consent

No informed consent was obtained because this study does not use human subjects or animals.

